# Prognostic impact and causality of age on oncological outcomes in women with endometrial cancer: a multimethod analysis of the randomised PORTEC-1, -2 and -3 trials

**DOI:** 10.1101/2023.10.31.23297837

**Authors:** Famke Wakkerman, Jiqing Wu, Hein Putter, Ina M. Jürgenliemk-Schulz, Jan J Jobsen, Ludy C.H.W. Lutgens, Marie A.D. Haverkort, Marianne de Jong, Jan Willem M. Mens, Bastiaan G. Wortman, Remi A. Nout, Alicia Léon-Castillo, Melanie E. Powell, Linda R. Mileshkin, Dionyssios Katsaros, Joanne Alfieri, Alexandra Leary, Naveena Singh, Stephanie M. de Boer, Hans W. Nijman, Vincent T.H.B.M. Smit, Tjalling Bosse, Viktor H. Koelzer, Carien L. Creutzberg, Nanda Horeweg

## Abstract

**Background:** Numerous studies have shown that elderly women with endometrial cancer (EC) have a higher risk of recurrence and cancer-related death. It is, however, unclear whether aging is a causal prognostic factor, or whether other risk factors become increasingly common with age. We address to this with a unique multi-method study design using state of the art statistical and causal inference techniques on datasets of three large randomised trials.

**Methods:** Data of 1801 women participating in the randomised PORTEC-1, -2 and -3 trials were used for statistical analyses and causal inference. The cohort included 714 patients with intermediate-risk EC, 427 high-intermediate risk EC patients and 660 high-risk EC patients. Associations of age with clinicopathological and molecular features were analysed using non-parametric tests. Multivariable competing risk analyses were performed to determine the independent prognostic value of age. To analyse age as a causal prognostic variable a deep learning Causal Inference model called AutoCI was used.

**Findings:** Median follow-up was 12·3 years for PORTEC-1, 10·5 years for PORTEC-2 and 6·1 years for PORTEC-3. Both overall recurrence and EC-specific deaths significantly increased with age. Moreover, elderly women had a higher incidence of deep myometrial invasion, serous tumour histology and p53abn tumours. Age was an independent risk factor for both overall recurrence (HR 1·02 per year, 95%CI 1·01-1·04; p=0·0012) and EC-specific death (HR 1·03 per year, 95%CI 1·01-1·05; p=0·0012), and was identified as a significant causal variable.

**Interpretation:** This study shows that advanced age is associated with more aggressive tumour features, and independently and causally related to worse oncological outcomes. Therefore, treatment for endometrial cancer in elderly women should not be de-escalated based on their age alone.

**Funding:** The PORTEC-1, -2 and -3 trials and the associated translational studies are supported by the Dutch Cancer Society.

## Introduction

Endometrial cancer (EC) is the sixth most common cancer in women worldwide, and its incidence is rising. This is in part due to ageing of the population, as EC is mostly a disease of postmenopausal women. For decades, it has been known that elderly women with EC have a worse prognosis than younger women. 70% of deaths from EC occur in women above 65 years of age.^1^ However, it remains unclear whether advanced age is an independent risk factor for cancer-specific death, or if other risk factors become increasingly common with older age. This may have contributed to the fact that age is not incorporated in most risk stratification systems for EC.

To assess the prognostic impact of age, only the relation with EC-specific outcomes is informative, as advanced age is causally related to a limited overall survival. Nonetheless, many studies only evaluate the relation of age with overall survival.^2^ Moreover, studies that do investigate recurrence or EC-specific survival often do not account for competing causes of death, which are more common in elderly, possibly leading to an overestimation of the impact of advanced age on prognosis.

The worse oncological outcomes in elderly women with EC could have several causes. One may be less intensive treatment. Due to comorbidities and frailty, older women are less likely to receive aggressive surgical treatment and adjuvant therapy.^3^ Moreover, age alone may be considered a reason for treatment de-escalation by some.^4^ Hence, results of studies where treatment is given at the discretion of the physician are potentially biased. Other causes of poorer oncological outcomes in elderly may be more aggressive tumour characteristics. Increasing age has been linked to a higher incidence of serous histology, deep myometrial invasion and high tumour grade.^4,5^

To determine whether advanced age is an independent risk factor, a comprehensive analysis correcting for all established clinicopathological and molecular risk factors is required. To date, some studies have corrected for the established clinicopathological risk factors,^6,7^ but the risk contribution of age has not been comprehensively investigated in the context of molecular classification, which has proven prognostic value.^5,8^

If age proves to be a significant independent risk factor, then quantification of the prognostic impact across age groups becomes relevant. Some studies have simplified the continuous nature of aging into categories or even dichotomized using a cut-off value. Often, a cut-off of 60 years is used,^9,10^ but no validation studies to support this arbitrary cut-off have been published.

Finally, all studies that have investigated the prognostic impact of age in EC so far are limited by their methodologies based on statistical correlation, rather than causation. Recently, causal inference using deep learning has been developed and tested on EC datasets.^11^ Under the assumption of a structural causal model^12^, this automated causal inference (AutoCI) technique identifies causal variables, which provides a causal re-interpretation that is supplementary to the standard statistical analysis.

Here, we address the question whether age is an independent risk factor for EC-specific outcomes in the context of clinicopathological and molecular characteristics, with a unique multi-method study design using state of the art statistical and causal inference techniques on datasets of three large randomised trials with a total study population of 1801 patients. We aim to determine how EC characteristics differ between younger and older women, and whether advanced age is independently and causally related to worse oncological outcomes.

## Methods

### Patient data

#### The randomised PORTEC-1, -2 and -3 trials

Data from the randomised PORTEC-1, -2 and -3 trials were used for statistical analyses and causal inference. Design and results of all three trials have been published.^13-15^ Briefly, in the PORTEC-1 trial, 714 women with early stage, intermediate risk endometrioid EC were randomly allocated to adjuvant external beam radiotherapy (EBRT) or no adjuvant therapy. Inclusion criteria were postoperative (FIGO 1988) stage I and either i) grade 1 with deep (>50%) myometrial invasion, or ii) grade 2, or iii) grade 3 with <50% myometrial invasion. The PORTEC-2 trial included 427 women with endometrioid EC with high-intermediate risk features, who were randomly allocated to either EBRT or vaginal brachytherapy (VBT). Inclusion criteria were (i) age >60 years and (FIGO 1988) stage IC and grade 1-2 disease, ii) age >60 years and stage IB, grade 3 disease, iii) any age with stage IIA (except grade 3 disease with >50% myometrial invasion). The PORTEC-3 trial recruited 660 women with high-risk stage I-III EC who were randomly allocated to chemoradiation or EBRT. The PORTEC-3 entry criteria were i) (FIGO 2009) stage IA, grade 3 endometrioid EC with lymphovascular space invasion (LVSI), ii) stage IB, grade 3 endometrioid EC, iii) endometrioid EC stage II-IIIC, iv) stage I-III EC with serous or clear cell histology. The study CONSORT diagram is provided in Supplementary Figure 1.

#### Pathology review and assessment of molecular class

In the PORTEC-1 and -2 trials, central pathology review was done after randomisation to assess stage (FIGO 1988), histotype, grade and LVSI. For both studies, LVSI was scored using a three-tiered quantification system (none vs. focal vs. substantial). In the PORTEC-3 trial, up-front central pathology review was performed to assess stage (FIGO 2009), histotype, grade and LVSI (two-tiered: absent vs. present). For the current study, all tumours were restaged according to FIGO 2009 and LVSI was dichotomised into absent (none and focal for PORTEC-1 and -2 patients) vs. present (substantial for PORTEC-1 and -2). Assignment of the molecular class of EC was performed according to the WHO 2020 classification of female genital tumours. More details are provided in Data Supplement 1.

#### Definition of endpoints

Primary endpoints for the current study were overall recurrence and EC-specific death. Secondary endpoints were distant metastasis, locoregional recurrence, pelvic recurrence and vaginal recurrence. Locoregional recurrence includes both vaginal and pelvic recurrence events.

### Methods statistical inference

Associations of age with clinicopathological and molecular features were analysed using non-parametric tests (the Mann-Whitney U and Kruskal Wallis tests).

The time-to-event was calculated from the date of randomisation to the date of recurrence, or to the date of death due to endometrial cancer. Patients who had no event were censored at the date of death due to other causes than EC, or date of last follow-up for those alive. For the outcome EC-specific death the competing event was death due to other causes than EC. For the outcomes overall recurrence and distant metastasis, the competing event was death due to any cause. For the outcomes locoregional, pelvic and vaginal recurrence, the competing events were both death due to any cause and distant metastasis.

Cause-specific cumulative incidence functions were calculated according to the Aalen-Johansen method, as described in Putter et al.^16^

Multivariable competing risk analyses were performed to determine the independent prognostic value of age as a continuous variable on the cause-specific hazard, corrected for the following pre-defined established risk factors: FIGO 2009 stage, histotype, grade, LVSI, molecular class and adjuvant treatment. For pelvic and vaginal recurrence, the correction for confounding was limited to stage, LVSI, molecular class and adjuvant treatment due to the lower number of events. In addition, all models were stratified by trial to correct for any cohort effects. Model fit was evaluated through time-dependent area under the receiver-operating characteristic curve (AUC) at 5-year follow-up and Brier scores for all outcomes.

Covariate data was missing in 510 patients (28.3%) for the molecular classification and in 185 patients (10.3%) for LVSI, and were imputed by multiple imputation using smcfcs (https://cran.r-project.org/package=smcfcs). To assess linearity of the effect of age, natural splines with 3 degrees of freedom were used. Statistical significance was accepted at p-values <0.01, always using two-sided tests.

### Methods causal inference

We utilised the Automated Causal Inference method as previously described^11^, to achieve an in-depth understanding of patient age as a causal prognostic variable for EC clinical outcomes. For this, a specific neural network architecture, i.e., the combination of a causal weight layer and a multilayer perceptron (MLP) is used for running the experiments. This model choice has shown optimal performance as compared to other previously published methodologies for the identification of causally-related variables with outcome in large clinical trial datasets including the PORTEC-1 and -2 cohorts.^11^

For analysis of the causal association of patient age with clinical outcomes, we grouped the data samples into three categories: <60 years (reference), 60-70 years, and >70 years. We then fed the age, clinicopathological, molecular, and adjuvant treatment variables to the neural network and supervised the model training with the Cox proportional hazards loss.^17^ Depending on how the hypothetical exclusion of a given variable would have impacted the maximum Frechet inception score on PORTEC-1, -2 and -3 data samples, we determined its causal association to the relevant EC outcome.^18^

To achieve reliable causal identification and hazard ratio (HR) computation, we conducted each experiment by repeatedly and randomly launching the training process 16 times for each endpoint of interest. Due to the non-linear nature of the neural network, we then used the (mean) gradient of input variables to compute the HR. Subsequently, the 95% confidence interval (CI) and p-value are reported based on the gradients collected from training the model 16 times. An in-depth explanation can be found in Data Supplement 2.

## Results

### Relation of age with clinicopathological and molecular variables of EC

All 1801 women of the intention-to-treat populations of the PORTEC-1, -2 and -3 trials were included (Table 1). Overall, their median age was 66 years (range 27-90 years). About one third (34.0%) of women included in PORTEC-1 were >70 years of age. In PORTEC-2, almost half of women (45.7%) were >70 years, while in the PORTEC-3 trial (which involved chemotherapy) only 18.2% of patients were >70 years. The relation of age with clinicopathological and molecular characteristics of EC is shown in Table 2.

**Table 1.** Patient and tumour characteristics by PORTEC-trial.

| Patient characteristics | PORTEC-1<br>n=714 | PORTEC-2<br>n=427 | PORTEC-3<br>n=660 | Total<br>n=1801 |
| --- | --- | --- | --- | --- |
| **Age at randomisation** | 66 (59-73) | 69 (65-75) | 62 (56-68) | 66 (60-72) |
| **Lymphadenectomy** |  |  |  |  |
| Not performed | 714 (100%) | 427 (100%) | 281 (42.6%) | 1422 (79.0%) |
| Performed | 0 (0%) | 0 (0%) | 379 (57.4%) | 379 (21.0%) |
| **Randomly allocated treatment** |  |  |  |  |
| None | 360 (50.4%)<sup>1</sup> | 0 (0%) | 0 (0%) | 360 (20.0%) |
| Brachytherapy | 0 (0%) | 213 (49.9%)<sup>2</sup> | 0 (0%) | 213 (11.8%) |
| External beam RT | 354 (49.6%) | 214 (50.1%) | 330 (50.0%)<sup>3,4</sup> | 898 (49.9%) |
| Chemoradiation | 0 (0%) | 0 (0%) | 330 (50.0%)<sup>5</sup> | 330 (18.3%) |
| **Stage (FIGO 2009)** |  |  |  |  |
| IA | 294 (41.2%) | 71 (16.6%) | 78 (11.8%) | 443 (24.6%) |
| IB | 420 (58.8%) | 351 (82.2%) | 117 (17.7%) | 888 (49.3%) |
| II | 0 (0%) | 2 (0.5%) | 170 (25.8%) | 172 (9.6%) |
| III | 0 (0%) | 3 (0.7%) | 295 (44.7%) | 298 (16.5%) |
| **Histograde** |  |  |  |  |
| Grade 1-2 EEC | 601 (84.2%) | 374 (87.6%) | 257 (38.9%) | 1232 (68.4%) |
| Grade 3 EEC | 95 (13.3%) | 41 (9.6%) | 185 (28.0%) | 321 (17.8%) |
| NEEC | 18 (2.5%) | 12 (2.8%) | 218 (33.0%) | 248 (13.8%) |
| **Lymphovascular space invasion** |  |  |  |  |
| Absent | 537 (75.2%) | 373 (87.4%) | 271 (41.4%) | 1181 (65.6%) |
| Present | 26 (3.6%) | 20 (4.7%) | 389 (58.9%) | 435 (24.2%) |
| Missing | 151 (8.4%) | 34 (8.0%) | 0 (0%) | 185 (10.3%) |
| **Molecular class** |  |  |  |  |
| *POLE*mut | 42 (5.9%) | 24 (5.6%) | 51 (7.7%) | 117 (6.5%) |
| MMRd | 137 (19.2%) | 110 (25.8%) | 139 (21.1%) | 386 (21.4%) |
| p53abn | 40 (5.6%) | 30 (7.0%) | 99 (15.0%) | 169 (9.4%) |
| NSMP | 265 (37.1%) | 232 (54.3%) | 122 (18.5%) | 619 (34.4%) |
| Missing | 230 (32.2%) | 31 (7.3%) | 249 (37.7%) | 510 (28.3%) |
| **ER status<sup>6</sup>** |  |  |  |  |
| Negative | 29 (4.1%) | 18 (4.2%) | 93 (14.1%) | 140 (7.8%) |
| Positive | 396 (55.5%) | 385 (90.2%) | 295 (44.7%) | 1076 (59.7%) |
| Missing | 289 (40.5%) | 24 (5.6%) | 272 (41.2%) | 585 (32.5%) |

Data are n (%), or median (IQR). Definition of abbreviations: RT = radiotherapy. EEC = endometrioid endometrial cancer. NEEC = non-endometrioid endometrial cancer. *POLE*mut = *POLE*-mutated.
MMRd = mismatch-repair deficient. p53abn = p53 abnormal. NSMP = no specific molecular profile.
1) For the PORTEC-1 trial, 369 patients received no adjuvant treatment and 345 patients received EBRT. 2) For the PORTEC-2 trial, 3 patients received no adjuvant treatment, 215 received brachytherapy and 209 received EBRT. 3) For the PORTEC-3 trial, 333 patients received EBRT and 327 patients received chemoradiation. 4) 158 patients received a vaginal brachytherapy boost, along their EBRT. 5) 151 patients received a vaginal brachytherapy boost, along their chemoradiation. 6) >10% nuclear ER expression was considered ER positive.

**Table 2.** Clinicopathological and molecular characteristics by age in the combined PORTEC-1, -2, -3 cohort.

| Age groups (years) | <60<br>n = 500 | 60-<70<br>n = 743 | ≥70<br>n = 558 | p value |
| --- | --- | --- | --- | --- |
| **Randomly allocated treatment** |  |  |  | **<0.0001<sup>2</sup>** |
| None | 107 (21.4%) | 135 (18.2%) | 118 (21.1%) |  |
| Brachytherapy | 16 (3.2%) | 93 (12.5%) | 104 (18.6%) |  |
| External beam RT | 249 (49.8%) | 371 (49.9%) | 278 (49.8%) |  |
| Chemoradiation | 128 (25.6%) | 144 (19.4%) | 58 (10.4%) |  |
| **Stage** |  |  |  | **<0.0001<sup>2</sup>** |
| IA | 150 (30.0%) | 172 (23.1%) | 121 (21.7%) |  |
| IB | 153 (30.6%) | 380 (51.1%) | 355 (63.6%) |  |
| II | 70 (14.0%) | 74 (10.0%) | 28 (5.0%) |  |
| III | 127 (25.4%) | 117 (15.7%) | 54 (9.7%) |  |
| **Histograde** |  |  |  | **0.001<sup>2</sup>** |
| Grade 1-2 EEC | 318 (63.6%) | 502 (67.6%) | 412 (73.8%) |  |
| Grade 3 EEC | 113 (22.6%) | 134 (18.0%) | 74 (13.3%) |  |
| SEC | 30 (6.0%) | 52 (7.0%) | 43 (7.7%) |  |
| CCC | 24 (4.8%) | 27 (3.6%) | 15 (2.7%) |  |
| Other<sup>7</sup> | 15 (3.0%) | 28 (3.8%) | 14 (2.5%) |  |
| **Myometrial invasion<sup>3</sup>** |  |  |  | **<0.0001<sup>1</sup>** |
| < 50% | 231 (46.4%) | 233 (31.4%) | 140 (25.1%) |  |
| ≥ 50% | 267 (53.6%) | 509 (68.6%) | 418 (74.9%) |  |
| **Lymphovascular space invasion<sup>4</sup>** |  |  |  | **<0.0001<sup>1</sup>** |
| None or focal<sup>8</sup> | 288 (64.6%) | 482 (71.9%) | 411 (82.2%) |  |
| Substantial or any | 158 (35.4%) | 188 (28.1%) | 89 (17.8%) |  |
| **Molecular class<sup>5</sup>** |  |  |  | **<0.0001<sup>2</sup>** |
| *POLE*mut | 63 (18.6%) | 32 (6.0%) | 22 (5.3%) |  |
| MMRd | 108 (31.9%) | 158 (29.5%) | 120 (28.8%) |  |
| p53abn | 23 (6.8%) | 82 (15.3%) | 64 (15.3%) |  |
| NSMP | 145 (42.8%) | 263 (49.2%) | 211 (50.6%) |  |
| **ER status<sup>6</sup>** |  |  |  | **0.788<sup>1</sup>** |
| Negative | 32 (10.8%) | 64 (12.5%) | 44 (10.8%) |  |
| Positive<sup>9</sup> | 265 (89.2%) | 448 (87.5%) | 363 (89.2%) |  |

Data are n (%). Definition of abbreviations: RT = radiotherapy. EEC = endometrioid endometrial cancer. SEC = serous endometrial cancer. CCC = clear cell endometrial cancer. *POLE*mut = *POLE* mutated. MMRd = mismatch-repair deficient. p53abn = p53 abnormal. NSMP = no specific molecular profile.
1) Mann-Whitney U test. 2) Kruskal-Wallis test. 3) 3 missing. 4) 185 missing. 5) 510 missing. 6) 585 missing. 7) Other: mixed endometrioid and clear cell EC, or mixed serous and clear cell EC, or mucinous adenocarcinoma, or carcinosarcoma, or undifferentiated. 8) For the PORTEC-1 and -2 trial, LVSI was scored using a three-tiered system (none, focal or substantial LVSI); for the PORTEC-3 trial LVSI was scored as none or any. For this analysis, LVSI was dichotomised into none or focal versus substantial or any. 9) >10% nuclear ER expression was considered ER positive.

Overall, elderly patients in these studies seemed to have less stage II and III disease (Table 2). But, 99.6% of women included in the PORTEC-1 and -2 trials had stage I disease (Supplemental Table 1-2), and no significant relation of age with stage was demonstrated in PORTEC-3 (p=0.11, Supplementary Table 3). However, deep myometrial invasion was significantly more frequent in older than in younger women (p<0.0001, Table 2).

No evident association of age with histology and tumour grade was found in the pooled analysis. Grade 3 EEC was not more common among elderly in PORTEC-1 and -2 (Supplementary Table 1-2). However, non-endometrioid histotypes were more common with increasing age in PORTEC-3, specifically serous EC (p<0.0001, Supplementary Table 3).

In the analysis of the three combined trials, substantial LVSI was found less often in elderly patients. LVSI was rare (4.0%) in PORTEC-1 and -2, and in the analyses of each trial separately, no significant relation of age with LVSI was found.

Analyses showed a significant difference in molecular classification between the different age groups (p<0.0001, Table 2). With rising age, there was a strong decrease in incidence of *POLE* mutated tumours, while incidence of p53abn tumours strongly increased. This difference was most striking between the group <60 years and the group 60-<70 years. The proportion of MMRd and NSMP tumours was stable across age groups (Table 2). Figure 1 illustrates the distribution of age for each molecular class.

**Figure 1.**
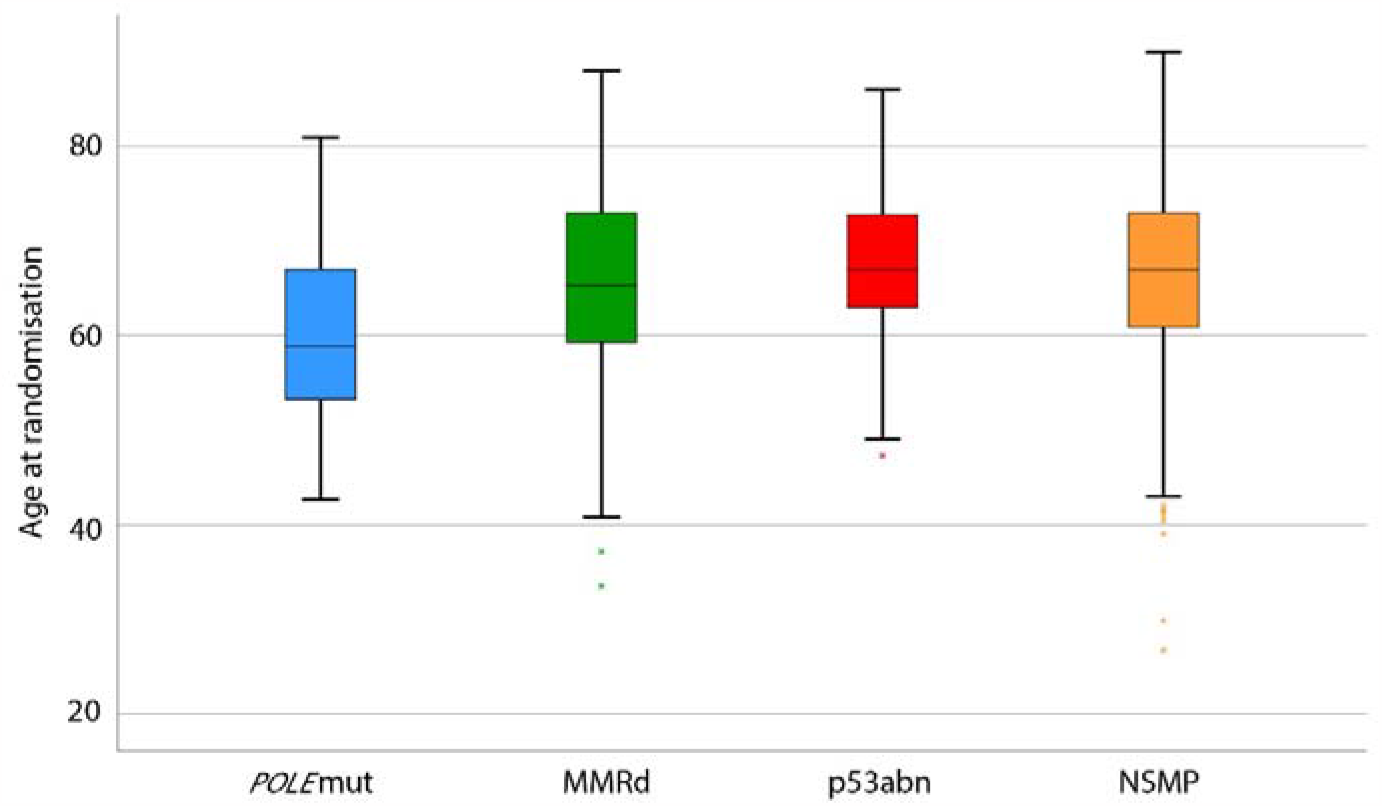
Distribution of age for each molecular class. Definition of abbreviations: *POLE*mut = POLE mutated. MMRd = mismatch-repair deficient. p53abn = p53 abnormal. NSMP = no specific molecular profile. Lowest line is minimum. Upper line is maximum. Box is first quartile, median and third quartile. Crosses are outliers.

No significant relation was observed between age and ER status in the combined cohorts. However, ER negative tumours were more common with increasing age in PORTEC-3 (p<0.0001, Supplementary Table 3).

#### Prognostic and causal impact of age

Patients were randomised 1:1 to adjuvant treatment in the PORTEC trials, and no differences in randomly allocated and received adjuvant treatment were observed between the age groups. Almost none of the patients in PORTEC-1 and -2 did not complete their treatment (0.6% for PORTEC-1 and 0.9% for PORTEC-2), with no differences between the age groups. In PORTEC-3, 52 patients (15.8%) did not complete chemotherapy (0-4 of the 6 cycles). Non-completion of chemotherapy was more common in elderly patients: 12.5% in women < 60 years, 16.0% in women 60-70 years old, and 22.4% in women > 70 years, however this was not significant (p=0.073). Median follow-up was 12.3 years (95%CI 11.9-12.6) in PORTEC-1, 10.5 years (95%CI 10.2-10.7) in PORTEC-2 and 6.1 years (95%CI 5.9-6.3) in PORTEC-3.

Both overall recurrence and EC-specific deaths significantly increased with age, with the largest differences observed between the age group <60 years and the other age groups (Figure 2). Cumulative incidence of all other oncological outcomes can be found in Supplementary Figure 2.

**Figure 2.**
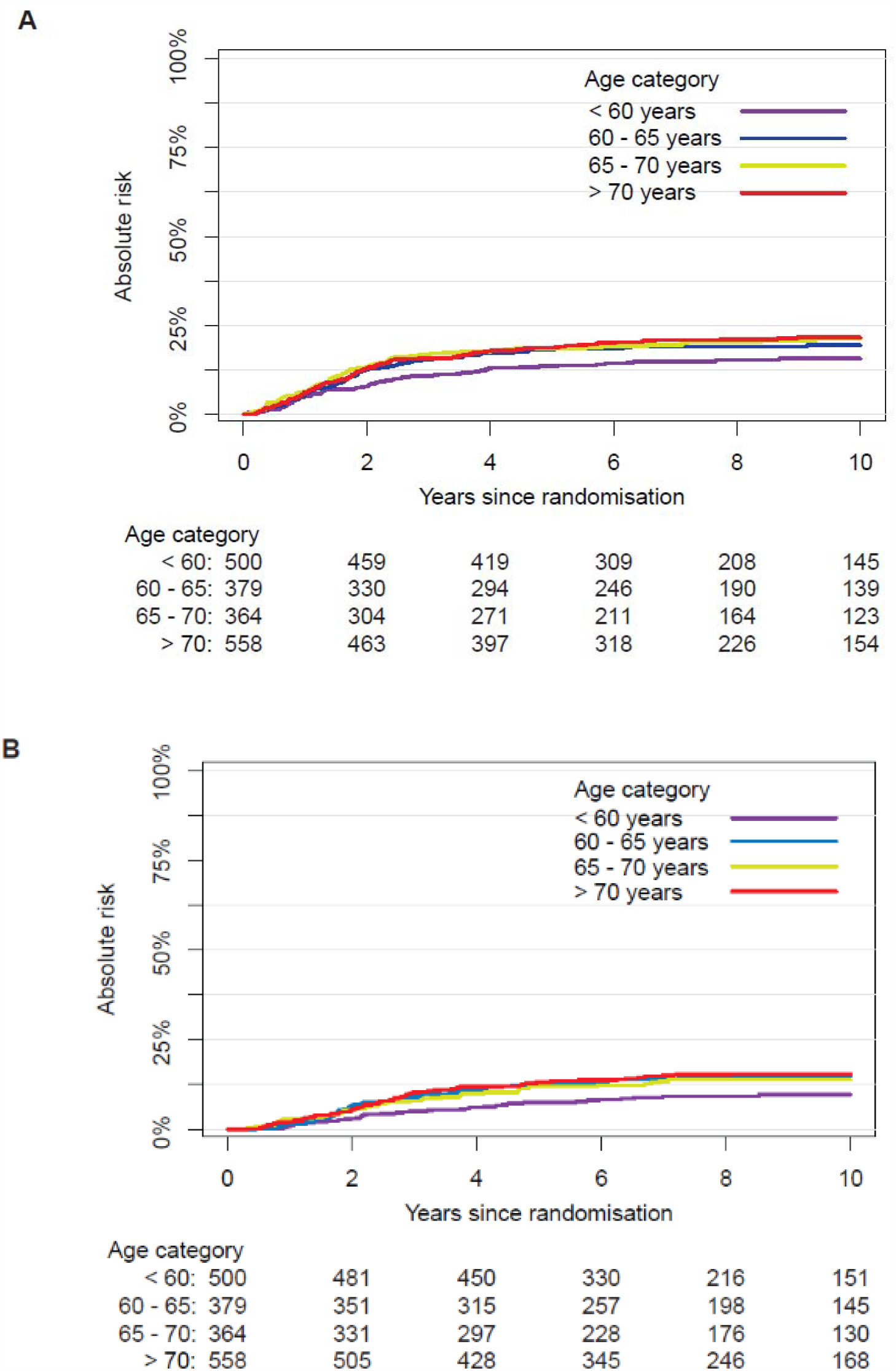
Cumulative incidence of overall recurrence and EC-specific death by age group. A) Cumulative incidence of overall recurrence by age group. B) Cumulative incidence of EC-specific death by age group.

Multivariable competing risk models were built for all outcomes and the AUCs ranged from 0.78 to 0.83, details and other performance metrics can be found in Supplementary Table 4. These multivariable competing risk analyses showed that age was an independent risk factor for overall recurrence with a cause-specific HR of 1.02 per year (95%CI 1.01-1.04; p=0.0012), corrected for stage, histotype and grade, LVSI, molecular class and adjuvant treatment. Likewise, age had an independent prognostic value for EC-specific death (HR 1.03, 95%CI 1.01-1.05; p=0.0012) and locoregional recurrence (HR 1.02, 95%CI 1.00-1.05; p=0.030). Age was also found to be an independent risk factor for vaginal recurrence (HR 1.05, 95%CI 1.01-1.08; p=0.0059), corrected for stage (I-II vs. III), LVSI, molecular class and adjuvant treatment (none vs. any type of adjuvant treatment). Age was not statistically associated with distant metastasis or pelvic recurrence.

The results above were supported by automated causal inference analyses, which identified age as a significant causal variable alongside all established clinicopathological and molecular risk factors for overall recurrence, EC-specific death and all other oncological outcomes (Table 3).

**Table 3.**
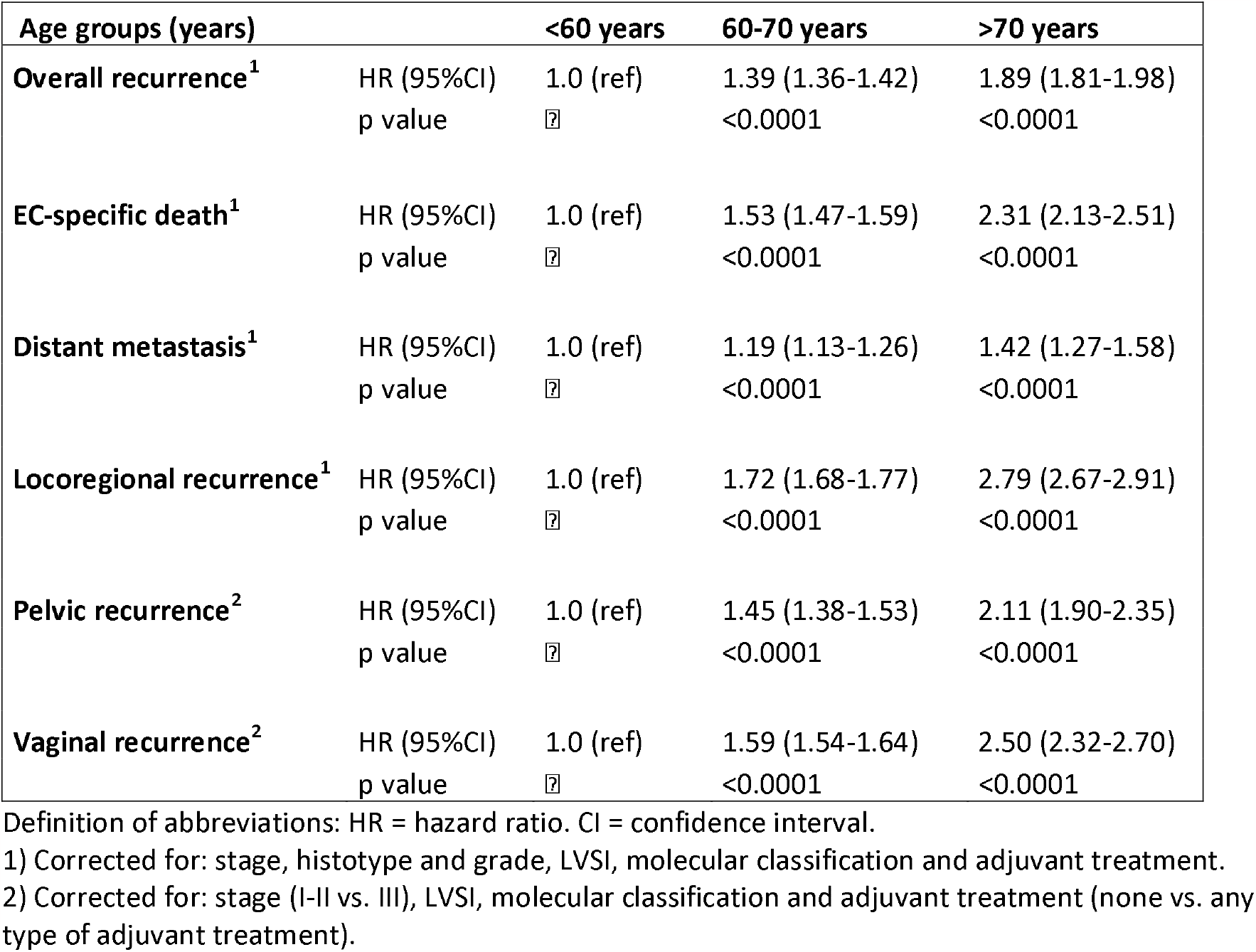
Prognostic impact of age using multivariable causal inference analysis.

#### Course of the risk increasing with age

Figure 3A and -B show the relation between age and the cause-specific hazard rates of overall recurrence and EC-specific death, as a hazard ratio, relative to age 60. The rates of both recurrence and EC-specific death increase steadily with an approximately linear course between the ages of 50 to 80 years (Figure 3). A similar relation of increased risk with age was observed for all secondary outcomes, although less strong (Supplementary Figure 3).

**Figure 3.**
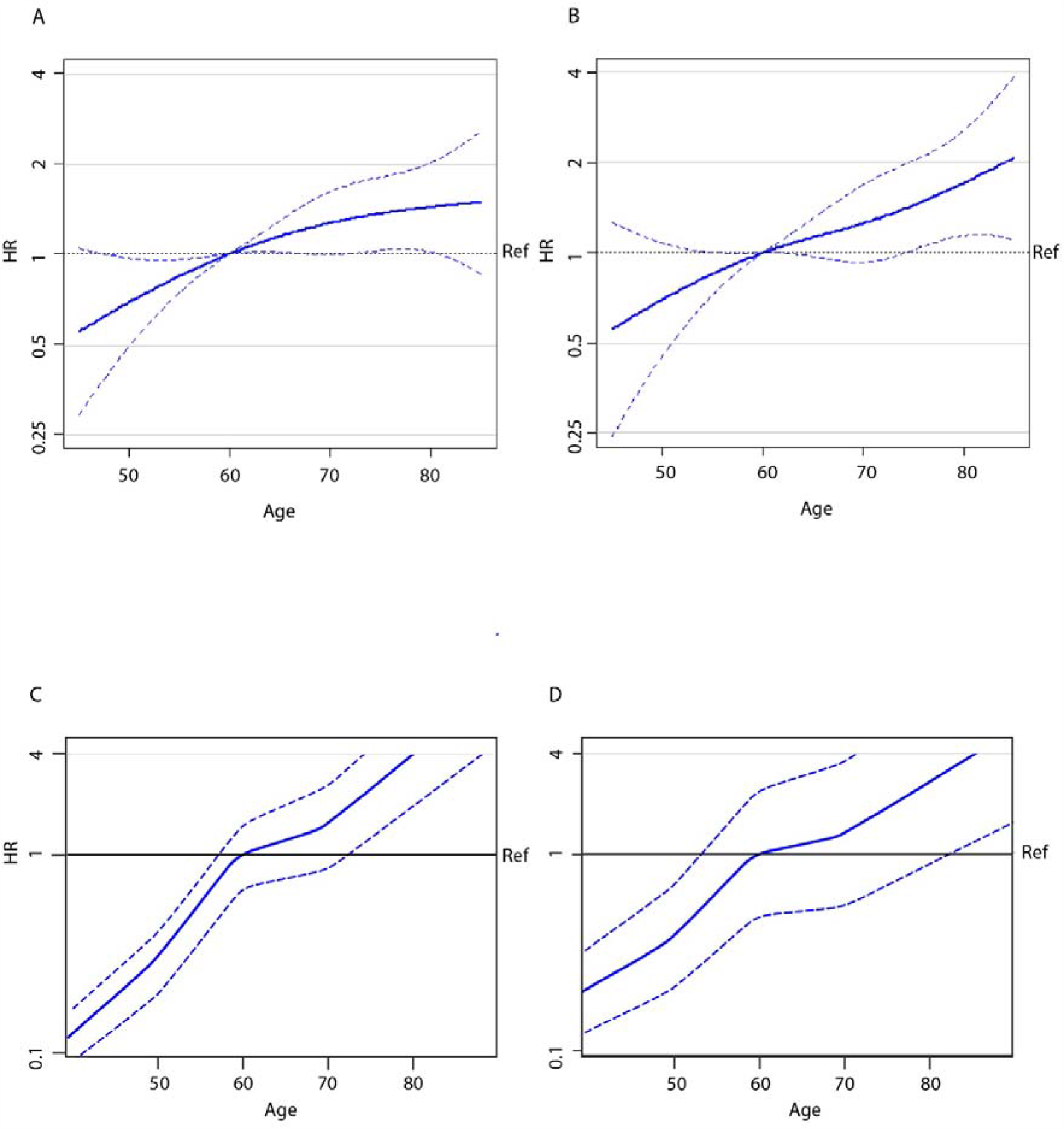
Relation of age and the rates of overall recurrence and EC-specific death. Definition of abbreviations: HR = hazard ratio. Ref = reference line. A) Correlation of age with the risk of overall recurrence by statistical inference. B) Correlation of age with the risk of EC-specific death by statistical inference. C) Correlation of age with the risk of overall recurrence by causal inference. D) Correlation of age with the risk of EC-specific death by causal inference.

In causal inference analysis, the correlation between age and both primary outcomes showed an accelerating increase of the cause-specific hazard rate with age tending towards an exponential relationship (Figure 3C-D). A similar pattern, though less strong, was observed for all secondary outcomes (Supplementary Figure 4).

## Discussion

In this pooled analysis of the randomised PORTEC-1, -2 and -3 trials, advanced age was associated with more aggressive tumour features, and independently and causally related to worse oncological outcomes. The risk of recurrence and EC-specific death steadily increased up to the age of 80.

This study confirmed that elderly women had a higher incidence of deep myometrial invasion, serous tumour histology and p53abn tumours, as seen by others.^2,4,5,7^ However, in this study the correlation between older age and more aggressive tumour characteristics does not entirely explain the poorer oncological outcomes in elderly women. Age was found to be an independent risk factor, after correcting for stage, histotype and grade, LVSI, molecular class, adjuvant treatment and competing events. Correction for molecular classification was done in two previous studies. Zheng et al.^5^ found age to be of independent prognostic value, after correcting for *TP53* mutation status and serous histology. Léon-Castillo et al.^8^ found age to be of independent prognostic value for recurrence-free survival when corrected for molecular group, in an analysis of the PORTEC-3 trial. The current study, using data of all three PORTEC-trials, is, to our knowledge, the first that investigates the prognostic impact of age across the whole spectrum of stage I-III EC, in the context of the molecular classification.

All studies published thus far have only analysed the correlation of age with prognosis using statistical inference. Awareness is increasing that many clinical questions that appear to be statistical associations at first glance, are essentially causal. This is due to the observation that conventional statistical analysis often falls short in generalising derived clinical claims for external validation.^19^ Causal inference aims to rigorously support the generalisability of experimental findings. Therefore, we applied a state-of-the-art causal inference method.^11^ To the best of our knowledge, the causal relation of age with oncological outcomes in women with endometrial cancer was investigated for the first time in this study. By using the cutting-edge AutoCI method that provides a novel linkage of causal inference and deep learning for the first time on three large randomised trials, we confirm that the relation of age with worse prognosis in endometrial cancer is causal.

Describing the change in risk of adverse oncological outcomes across different age groups is important for translation of these results to clinical practice. Our analyses showed that, the probability of adverse oncological outcomes increases steadily between the ages of 50 to 80 years, after correction for all established risk factors (Figure 3). We also performed a systematic literature search (See Research in Context) and found that only Sun et al.^20^ published a modelling study on the course of the increasing risk with age in EC. They found a similar linear increase of risk with age, with no clear cut-off value. However, the potential for clinical translation of their results is limited, as no correction for confounding was performed. Other studies that investigated the age to risk relation without proper survival analysis, did establish cut-off values of 63 and 65 years.^20,21^ Thus, the risk of adverse outcomes appears to keep increasing with age, and our systematic literature search showed no consensus on a cut-off value in the literature (Research in Context). This is supported by the fact that we found consistent HRs for the risk of adverse outcomes with increasing age in multiple studies that analysed different risk groups.^8,22^ All reported HRs were in the range of 2-4% per year, for both high-intermediate risk groups and high-risk groups.

Based on these results, it seems arbitrary to decide on a cut-off for age. This complicates the use of age as a risk factor, as risk tables in treatment guidelines are created using categorial variables. This may force guideline committees in the future to dichotomise age or decide on age groups regardless, or to even leave this significant risk factor out. Nonetheless, our results are of value for clinical practice, as they suggest that advanced age alone should not be a reason for treatment de-escalation. In clinical practice, comprehensive diagnostic assessment of all patients, young and old, is necessary. Assessment of the extent of disease and molecular classification of the tumour is essential to estimate risk of recurrence and death and guide treatment decisions,^8,23^ also in elderly women. Geriatric assessments can be useful to better determine individual risks of morbidity and mortality associated with adjuvant treatment. The results of these two types of assessments and the preferences of the elderly patient should be considered in a shared decision-making process on adjuvant therapy.

Strengths of this study are the use of three large randomised trials with high-quality long-term follow-up data and large number of molecularly classified tumours. The use of a multimodal analysis, based on state-of-the-art statistical and causal inference techniques is unique and identified age as a causal risk factor. Our study design ensured minimal bias, sufficient statistical power and high internal validity. The downside of using trial data is that participants are not representative of all patients in medical practice. Exclusion criteria for the PORTEC-trials included certain comorbidities, like impaired renal or cardiac function, which are more prevalent in elderly patients.^13-15^ Moreover, patients in the PORTEC-3 trial had to be fit enough to undergo chemoradiation. However, we do not expect that the prognostic impact of age would be different in a real-life population. The proportion of elderly women who are fit enough and willing to undergo surgery and adjuvant treatment will be smaller though, which affects risk of recurrence and death. Therefore, it is probable that the impact of age on prognosis is underestimated in this study. Another limitation of this study was the limited number of patients younger than 50 years and older than 80 years, subgroups for which we prefer not to draw any conclusions.

The pathophysiological mechanisms that explain the increased risk for elderly with EC are still unclear. Obtaining insight into the relationship between calendar age and biological age, and the ways in which cancer control is different in elderly, is of value as it may yield prognostic refinement or identify more appropriate therapies. Recent research has shown a correlation between increasing age and genomic instability of tumours. Chatsirisupachai et al. found that age-related genomic differences in tumours were largest in EC.^24^ These findings could be of interest in further research. Furthermore, we hypothesize that one factor influencing the worse prognosis of elderly is the decline of the immune system and anti-tumour response.^25,26^ Studies into the prognostic impact of anti-tumour response have revealed that the presence of tumour-infiltrating lymphocytes (TIL) or tertiary lymphoid structures are associated with a better prognosis.^27-29^ In breast cancer patients,^30^ younger patients have a significantly higher TIL density than older patients, which may explain in part why elderly women have a worse prognosis. These findings encourage further exploration of this topic in endometrial cancer specifically.

In conclusion, our study showed that elderly women with endometrial cancer have worse oncological outcomes, even when they undergo the same treatment as younger women. This is in part explained by more aggressive histopathological and molecular tumour features with advancing age, but an independent and causal negative effect of age remains unexplained. This risk for adverse oncological outcomes associated with aging gradually increases between 50 and 80 years without a clear cut-off. Therefore, elderly women with endometrial cancer should not be excluded from diagnostic assessments, molecular classification, and adjuvant therapy based on their age alone.

### Panel: Research in context

#### Systematic review

To create an overview of the current knowledge, a systematic literature search was performed to identify original studies investigating the impact of calendar age by multivariable analysis on oncological outcomes in women with EC. A search string based on Mesh terms and text words for the following elements was built: Endometrial cancer AND age AND risk factors AND survival analysis. The search was performed on April 13^th^ 2022. The search string and article selection and data extraction methods are detailed in Data Supplement 3 and 4.

#### Interpretation

A total of 783 articles were found, of which 108 were selected. Of those articles, 5 were from randomised controlled trials and 103 from prospective and retrospective cohort studies. As investigating the prognostic impact of age was not the primary objective of most studies, differences between the younger and older women in tumour characteristics and treatment were commonly observed. As a result, the question whether EC is intrinsically more aggressive in elderly patients has so far remained unanswered. Only one of the 108 included studies examined the risk of age in the context of the EC molecular classification, which has proven prognostic impact. No studies were found that investigated women of all risk groups of endometrial cancer in the context of the molecular classification. Lastly, little consensus was observed between studies on the use of any cut-offs for age (Supplemental Table 5). About a third of the studies analysed age as a continuous variable (n=35, 35.4%), another third chose a cut-off value of 60 years (n=33, 33.3%) and the remaining studies chose a variety of cut-offs. A few studies have modelled the age-risk relation and proposed different cut-offs at 55, 61, 63 and 65 years. These results make it questionable whether there is a single value for which the tipping point of increased risk is best represented, or whether the increased risk of age should be regarded as a continuous phenomenon, which may be simplified by subdivision into groups of, for example, 10 years. Thus, a comprehensive study of the relation of age with tumour characteristics and oncological outcomes in EC is warranted.

## Data Availability

The datasets used in this study are not publicly available due to restrictions by privacy laws. Data from PORTEC-1 and PORTEC-2 trial are held by the PORTEC study group and data from the PORTEC-3 trial are available to members of the international TransPORTEC Research Consortium. Requests for sharing of all data and material should be addressed to the corresponding author within 15 years of the date of publication of this Article and include a scientific proposal. Depending on the specific research proposal, the TransPORTEC consortium (PORTEC-3 study) or the PORTEC study group (PORTEC-1, PORTEC-2), will determine when, for how long, for which specific purposes, and under which conditions the requested data can be made available, subject to ethical consent.

