## Supplement for "Prognostic impact and causality of age on oncological outcomes in women with endometrial cancer: a multimethod analysis of the randomised PORTEC-1, -2 and -3 trials"

**Wakkerman F.C. et al.**

**Supplementary appendix**

### Supplementary figure 1. CONSORT flowchart PORTEC -1, -2 and -3

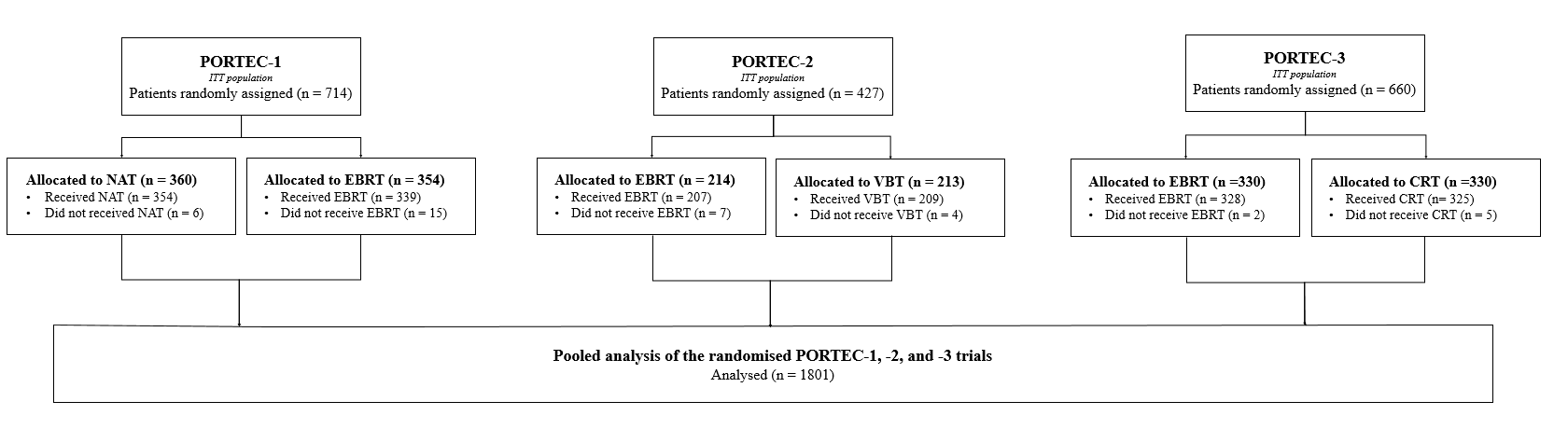

### Data Supplement 1. Assessment of molecular class

All cases with available tumour material were assessed for pathogenic variants in the exonuclease domain of DNA polymerase-ε (POLEmut), by Sanger sequencing in PORTEC-1 and -2 and next-generation sequencing in PORTEC-3.^1, 2^ Any tumour with one of the eleven established pathogenic variants was considered ‘POLEmut’ EC.^3^ Tumours that were POLE-wildtype, were classified as MMRd if immunohistochemical staining (IHC) for MMR proteins (MLH1, PMS2, MSH2, and MSH6) showed loss of nuclear staining in >10% of the tumour cells.^1, 4, 5^ If MMR IHC was not evaluable (failed or ambiguous cases), microsatellite instability (MSI) was assessed using the MSI analysis system, version 1.2 (Promega, Madison, WI). POLE wild type, MMR-proficient tumours were classified as p53abn if IHC showed mutant overexpression, or a null mutant, or cytoplasmic pattern. If p53 immunohistochemistry was ambiguous or had failed, TP53 mutational status was assessed by NGS.^6^ Tumours that were POLE wildtype, MMR proficient and p53 wildtype were classified as no specific molecular profile ‘NSMP EC’.^7, 8^

IHC staining for ER was performed on whole slides and was considered positive if more than 10% of the tumour showed positive nuclear staining. This cut-off is most commonly used for the assessment of ER expression in EC cases.^9-11^

### Data Supplement 2. Methods AutoCI

In a recent causal inference study^12^, researchers have demonstrated that AutoCI can be used to correctly quantify the causal likelihood of variables of interest for PORTEC -1 and -2. Importantly, the identification performance remains consistent in the presence of hidden confounders. Motivated by these observations, it is therefore natural to adapt the existing AutoCI model to the PORTEC -1, -2 and -3 cohorts, with the emphasis on the investigation of the age variable.

Instead of treating the age as the binary variable in^12^, here we split the patients into three age groups, e.g., <60 years (baseline), 60-70 years, >70 years, so we can carry out a comparable causal analysis to the statistical counterpart. To depict the continuous HR trend, we also take the raw age value as another variant of input formality for training the model (See Figure 2 in manuscript). For the outcomes, overall recurrence, EC-specific death, distant metastasis and locoregional recurrence, we further include stage, histotype and grade, LVSI, molecular group and adjuvant treatment as input variables. For the outcomes, pelvic and vaginal recurrence, we take stage, LVSI, molecular group and adjuvant treatment as input variables.

In the existing AutoCI paper^12^, ablative studies on (type-safe) neural architecture search have been conducted and the model that combines a causal weight layer and a multilayer perceptron (MLP) was demonstrated to be sufficient and successful in learning meaningful causal associations that link to the EC recurrence. Based on these findings, we directly utilise this optimal model and customise it to three PORTEC cohorts in this study.

Thanks to the differentiable property of Cox proportional hazards introduced in Katzman et al.^13^ and the implementation instantiated in the pycox package, we train the neural network with the Cox proportional hazards loss. Similar to the previous AutoCI paper^12^, we then apply the maximum Frechet inception score (FID), that is, the maximum of FIDs computed on PORTEC 1-3 data batches individually to determine the influence coming from the hypothetical exclusion of a given variable. Provided that the maximum FID increases significantly after the age variable is missing, we confirm that the age variable is causally related to oncological outcome(s). Following the recommendations suggested in^12^, the whole training process takes 8 epoches in the warm-up stage and then another 8 epochs for filtering out non-causal variables. In the meantime, we apply the popular Adam optimiser for updating the learnable weights during the training session and use the default hyper-parameter configurations.

To improve the reproducibility and reliability of model performance, all experiments are repeated 16 times and the collection of gradients with respect to each input variable are recorded as meaningful proxies for HR computation. Such a practice is driven by the non-linearity property of modern neural network in contrast to the common linearity assumption required by the Cox model. With the collection of gradients in hand, we report the mean HR together with the 95% CI and p-value (See Table 3 in manuscript).

In summary, our codes are implemented with the Python language and built upon the Python packages including pytorch, numpy, pandas, scipy and pycox. For more implementation details we refer interested readers to [https://github.com/CTPLab/AutoCI](about:blank) and [https://github.com/havakv/pycox](about:blank) .

Taking both the causal and statistical analysis as a whole, we provide a comprehensive and in-depth investigation on the (causal) role of age variable played in a variety of EC outcomes.

| **Age groups (years)** | **<60** | **60-<70** | **≥70** | **p value** |
| --- | --- | --- | --- | --- |
|  | **n=200** | **n=271** | **n=243** |  |
| **Randomly allocated treatment** | |  |  | **0.25^1^** |
| None | 107 (53.5%) | 135 (49.8%) | 118 (48.6%) |  |
| External beam RT | 93 (46.5%) | 136 (50.2%) | 125 (51.4%) |  |
| **Stage (FIGO 2009)** |  |  |  | **<0.0001^1^** |
| IA | 107 (53.5%) | 104 (38.4%) | 83 (34.2%) |  |
| IB | 93 (46.5%) | 167 (61.6%) | 160 (65.8%) |  |
| **Histograde** |  |  |  | **0.82^2^** |
| Grade 1-2 EEC | 167 (83.5%) | 228 (84.1%) | 206 (84.8%) |  |
| Grade 3 EEC | 28 (14.0%) | 37 (13.7%) | 30 (12.3%) |  |
| NEEC | 5 (2.5%) | 6 (2.2%) | 7 (2.9%) |  |
| **Lymphovascular space invasion^3^** | |  |  | **0.90^1^** |
| None or focal | 140 (95.2%) | 203 (94.9%) | 194 (96.0%) |  |
| Substantial | 7 (4.8%) | 11 (5.1%) | 8 (4.0%) |  |
| **Molecular class^4^** |  |  |  | **<0.0001^2^** |
| *POLE*mut | 26 (19.8%) | 9 (4.9%) | 7 (4.1%) |  |
| MMRd | 32 (24.4%) | 53 (29.1%) | 52 (30.4%) |  |
| p53abn | 6 (4.6%) | 14 (7.7%) | 20 (11.7%) |  |
| NSMP | 67 (51.1%) | 106 (58.2%) | 92 (53.8%) |  |
| **ER status^5^** |  |  |  |  |
| Negative | 8 (7.3%) | 10 (6.2%) | 11 (7.1%) | **0.98^1^** |
| Positive^6^ | 102 (92.7%) | 151 (93.8%) | 143 (92.9%) |  |

### Supplementary Table 1. Clinicopathological and molecular characteristics by age in the PORTEC-1 trial

Data are n (%). Definition of abbreviations: RT = radiotherapy. EEC = endometrioid endometrial cancer. NEEC = non-endometrioid endometrial cancer. *POLE*mut = *POLE*-ultramutated. MMRd = mismatch-repair deficient. p53abn = p53 abnormal. NSMP = no specific molecular profile.

1) Mann-Whitney U test. 2) Kruskal-Wallis test. 3) 151 missing. 4) 230 missing. 5) 289 missing. 6) >10% nuclear ER expression was considered ER positive.

### Supplementary Table 2. Clinicopathological and molecular characteristics by age in the PORTEC-2 trial

| **Age groups (years)** | **<60** | **60-<70** | **≥70** | **p value** |
| --- | --- | --- | --- | --- |
|  | **n=32** | **n=200** | **n=195** |  |
| **Randomly allocated treatment** |  |  |  | **0.38^1^** |
| External beam RT | 16 (50.0%) | 107 (53.5%) | 91 (46.7%) |  |
| Brachytherapy | 16 (50.0%) | 93 (46.5%) | 104 (53.3%) |  |
| **Stage (FIGO 2009)** |  |  |  | **0.005^2^** |
| IA | 18 (56.3%) | 31 (15.5%) | 22 (11.3%) |  |
| IB | 13 (40.6%) | 168 (84.0%) | 170 (87.2%) |  |
| II | 0 (0%) | 1 (0.5%) | 1 (0.5%) |  |
| III | 1 (3.1%) | 0 (0%) | 2 (1.0%) |  |
| **Histograde** |  |  |  | **0.25^2^** |
| Grade 1-2 EEC | 29 (90.6%) | 173 (86.5%) | 172 (88.2%) |  |
| Grade 3 EEC | 2 (6.3%) | 24 (12.0%) | 15 (7.7%) |  |
| NEEC | 1 (3.1%) | 3 (1.5%) | 8 (4.1%) |  |
| **Myometrial invasion** |  |  |  | **<0.0001^1^** |
| < 50% | 19 (59.4%) | 31 (15.5%) | 21 (10.8%) |  |
| ≥ 50% | 13 (40.6) | 169 (84.5%) | 174 (89.2%) |  |
| **Lymphovascular space invasion^3^** | |  |  | **0.68^1^** |
| None or focal | 28 (90.3%) | 175 (95.1%) | 170 (95.5%) |  |
| Substantial | 3 (9.7%) | 9 (4.9%) | 8 (4.5%) |  |
| **Molecular class^4^** |  |  |  | **0.80^2^** |
| *POLE*mut | 0 (0%) | 12 (6.5%) | 12 (6.6%) |  |
| MMRd | 10 (33.3%) | 47 (25.5%) | 53 (29.1%) |  |
| p53abn | 1 (3.3%) | 14 (7.6%) | 15 (8.2%) |  |
| NSMP | 19 (63.3%) | 111 (60.3%) | 102 (56.0%) |  |
| **ER status^5^** |  |  |  | **0.14^1^** |
| Negative | 1 (3.7%) | 6 (3.2%) | 11 (5.9%) |  |
| Positive^6^ | 26 (96.3%) | 182 (96.8%) | 177 (94.1%) |  |

Data are n (%). Definition of abbreviations: RT = radiotherapy. EEC = endometrioid endometrial cancer. NEEC = non-endometrioid endometrial cancer. POLEmut = POLE-ultramutated. MMRd = mismatch-repair deficient. p53abn = p53 abnormal. NSMP = no specific molecular profile.

1) Mann-Whitney U test. 2) Kruskal-Wallis test. 3) 34 missing. 4) 31 missing. 5) 24 missing. 6) >10% nuclear ER expression was considered ER positive.

| **Age groups (years)** | **<60** | **60-<70** | **≥70** | **p value** |
| --- | --- | --- | --- | --- |
|  | **n=268** | **n=272** | **n=120** |  |
| **Lymphadenectomy** |  |  |  | **0.62^1^** |
| Not performed | 116 (43.3%) | 111 (40.8%) | 54 (45.0%) |  |
| Performed | 152 (56.7%) | 161 (59.2%) | 66 (55.0%) |  |
| **Randomly allocated treatment** | |  |  | **0.68^1^** |
| External beam RT | 140 (52.2%) | 128 (47.1%) | 62 (51.7%) |  |
| Chemoradiation | 128 (47.8%) | 144 (52.9%) | 58 (48.3%) |  |
| **Stage (FIGO 2009)** |  |  |  | **0.11^2^** |
| IA | 25 (9.3%) | 37 (13.6%) | 16 (13.3%) |  |
| IB | 47 (17.5%) | 45 (16.5%) | 25 (20.8%) |  |
| II | 70 (26.1%) | 73 (26.8%) | 27 (22.5%) |  |
| III | 126 (47.0%) | 117 (43.0%) | 52 (43.3%) |  |
| **Histograde** |  |  |  | **<0.0001^2^** |
| Grade 1-2 EEC | 122 (45.5%) | 101 (37.1%) | 34 (28.3%) |  |
| Grade 3 EEC | 83 (31.0%) | 73 (26.8%) | 29 (24.2%) |  |
| SEC | 25 (9.3%) | 48 (17.6%) | 32 (26.7%) |  |
| CCC | 23 (8.6%) | 26 (9.6%) | 13 (10.8%) |  |
| Other^6^ | 15 (5.6%) | 24 (8.8%) | 12 (10.0%) |  |
| **Myometrial invasion^3^** |  |  |  | **0.07^1^** |
| < 50% | 105 (39.5%) | 98 (36.2%) | 36 (30.0%) |  |
| ≥ 50% | 161 (60.5%) | 173 (63.8%) | 84 (70.0%) |  |
| **Lymphovascular space invasion** | |  |  | **0.045^1^** |
| None | 120 (44.8%) | 104 (38.2%) | 47 (39.2%) |  |
| Any | 148 (55.2%) | 168 (61.8%) | 73 (60.8%) |  |
| **Molecular class^4^** |  |  |  | **<0.0001^2^** |
| *POLE*mut | 37 (20.8%) | 11 (6.5%) | 3 (4.7%) |  |
| MMRd | 66 (37.1%) | 58 (34.3%) | 15 (23.4%) |  |
| p53abn | 16 (9.0%) | 54 (32.0%) | 29 (45.3%) |  |
| NSMP | 59 (33.1%) | 46 (27.2%) | 17 (26.6%) |  |
| **ER status^5^** |  |  |  | **<0.0001^1^** |
| Negative | 23 (14.4%) | 48 (29.4%) | 22 (33.8%) |  |
| Positive^6^ | 137 (85.6%) | 115 (70.6%) | 43 (66.2%) |  |

### Supplementary Table 3. Clinicopathological and molecular characteristics by age in the PORTEC-3 trial

Data are n (%). Definition of abbreviations: RT = radiotherapy. EEC = endometrioid endometrial cancer. SEC = serous endometrial cancer. CCC = clear cell endometrial cancer. *POLE*mut = POLE-ultramutated. MMRd = mismatch-repair deficient. p53abn = p53 abnormal. NSMP = no specific molecular profile.

1) Mann-Whitney U test. 2) Kruskal-Wallis test. 3) 3 missing. 4) 249 missing. 5) 272 missing. 6) Other: mixed endometrioid and clear cell EC, or mixed serous and clear cell EC, or mucinous adenocarcinoma, or carcinosarcoma, or undifferentiated. 6) >10% nuclear ER expression was considered ER positive.

### Supplementary figure 2. Cumulative incidence by age group for secondary outcomes

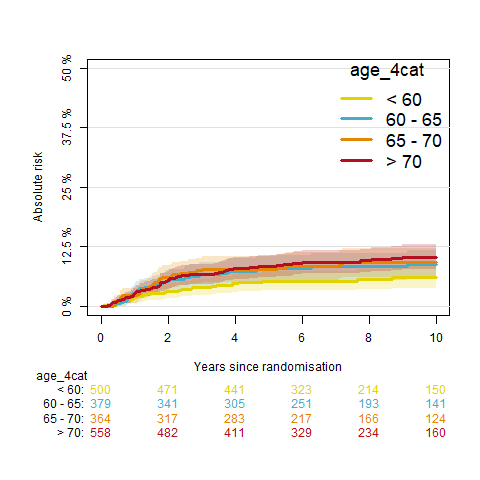

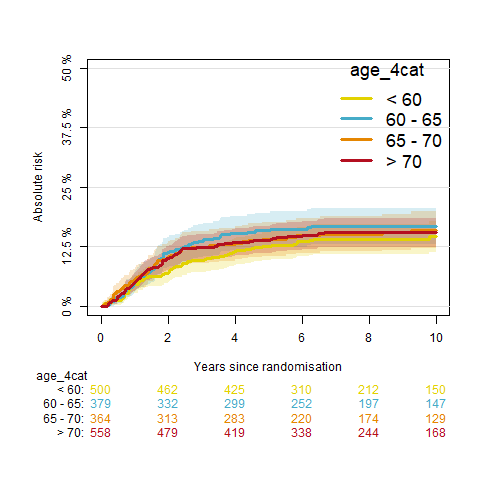
**A	B**

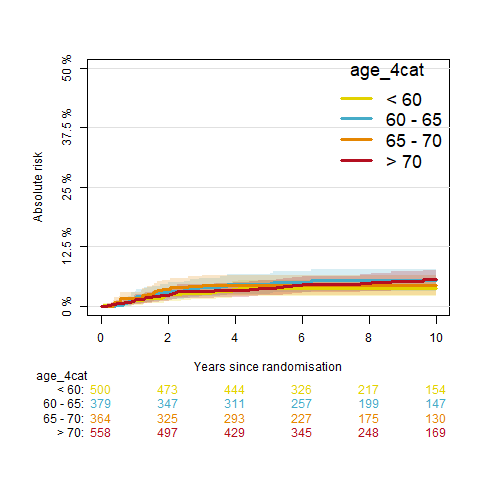
**C	D**

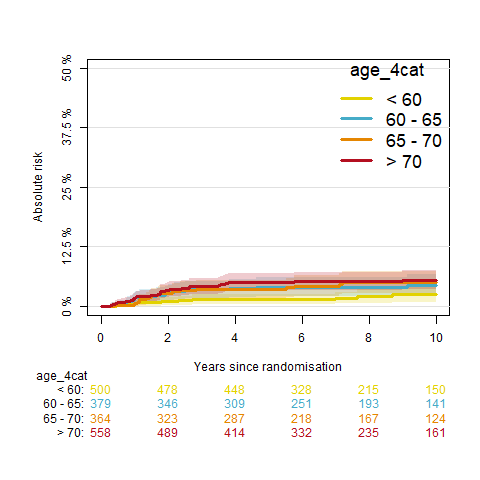

A) Cumulative incidence of distant metastasis by age group. B) Cumulative incidence of locoregional recurrence by age group. C) Cumulative incidence of pelvic recurrence by age group. D) Cumulative incidence of vaginal recurrence by age group.

### Supplementary Table 4. Time-dependent AUC and Brier score for all outcomes

| **Outcome** | **AUC (95%CI)^1^** | **Brier score (95%CI)** |
| --- | --- | --- |
| **Overall recurrence** | 0·78 (0·75-0·81) | 0·12 (0·11-0·13) |
| **EC-specific death** | 0·82 (0·78-0·85) | 0·08 (0·07-0·09) |
| **Distant metastasis** | 0·83 (0·80-0·86) | 0·09 (0·08-0·10) |
| **Locoregional recurrence** | 0·78 (0·74-0·82) | 0·06 (0·05-0·07) |
| **Pelvic recurrence** | 0·83 (0·78-0·88) | 0·04 (0·03-0·05) |
| **Vaginal recurrence** | 0·78 (0·73-0·84) | 0·03 (0·03-0·04) |
| Definition of abbreviations: AUC = area under receiver-operating characteristic curve. 1) Calculated at 5-year follow-up | | |

### Supplementary Figure 3. Correlation of age with the risk of secondary outcomes by statistical inference

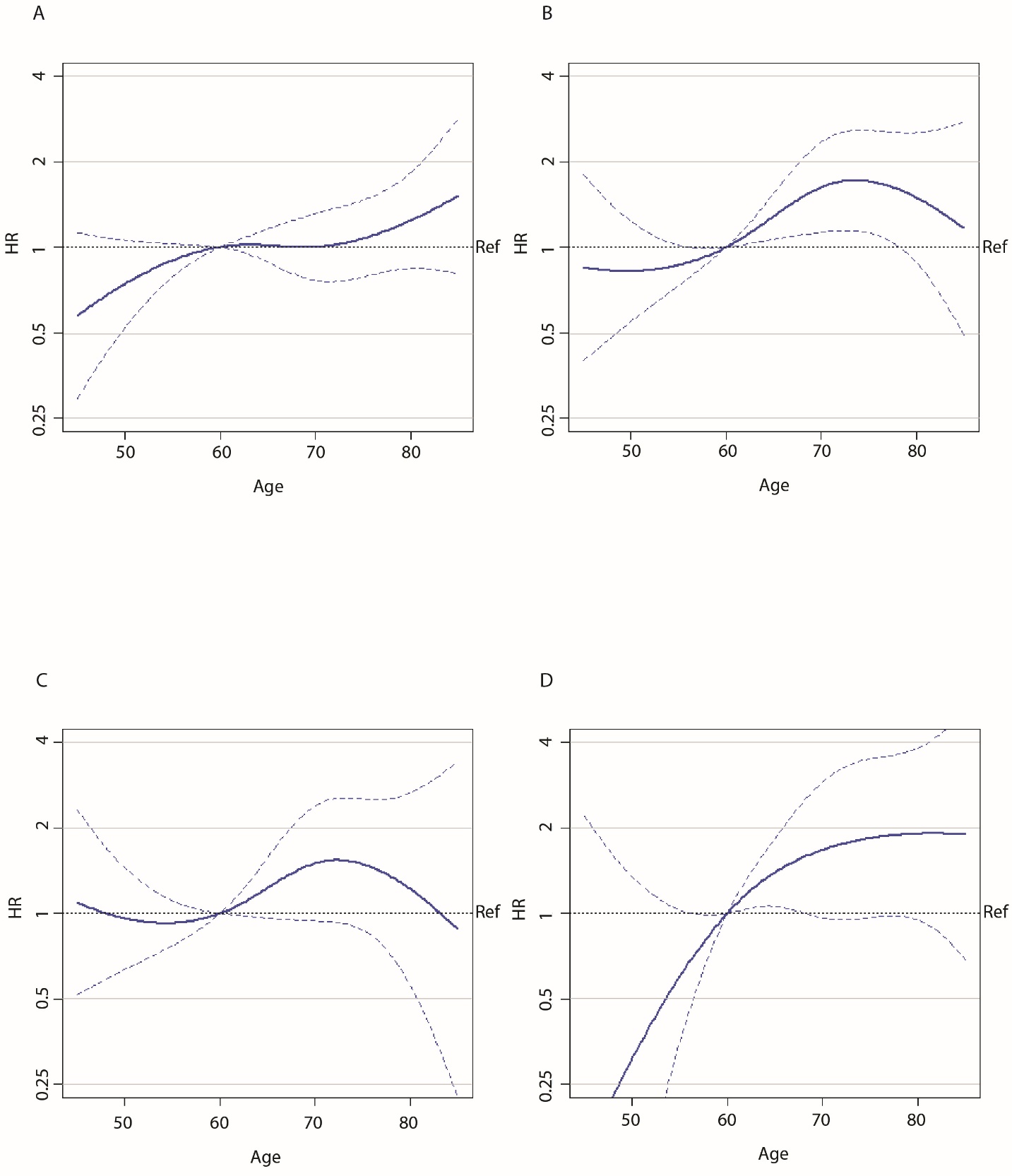

A) Correlation of age with the risk of distant metastasis by statistical inference. B) Correlation of age with the risk of locoregional recurrence by statistical inference. C) Correlation of age with the risk of pelvic recurrence by statistical inference. D) Correlation of age with the risk of vaginal recurrence by statistical inference. Definition of abbreviations: HR = hazard ratio. Ref = reference line.

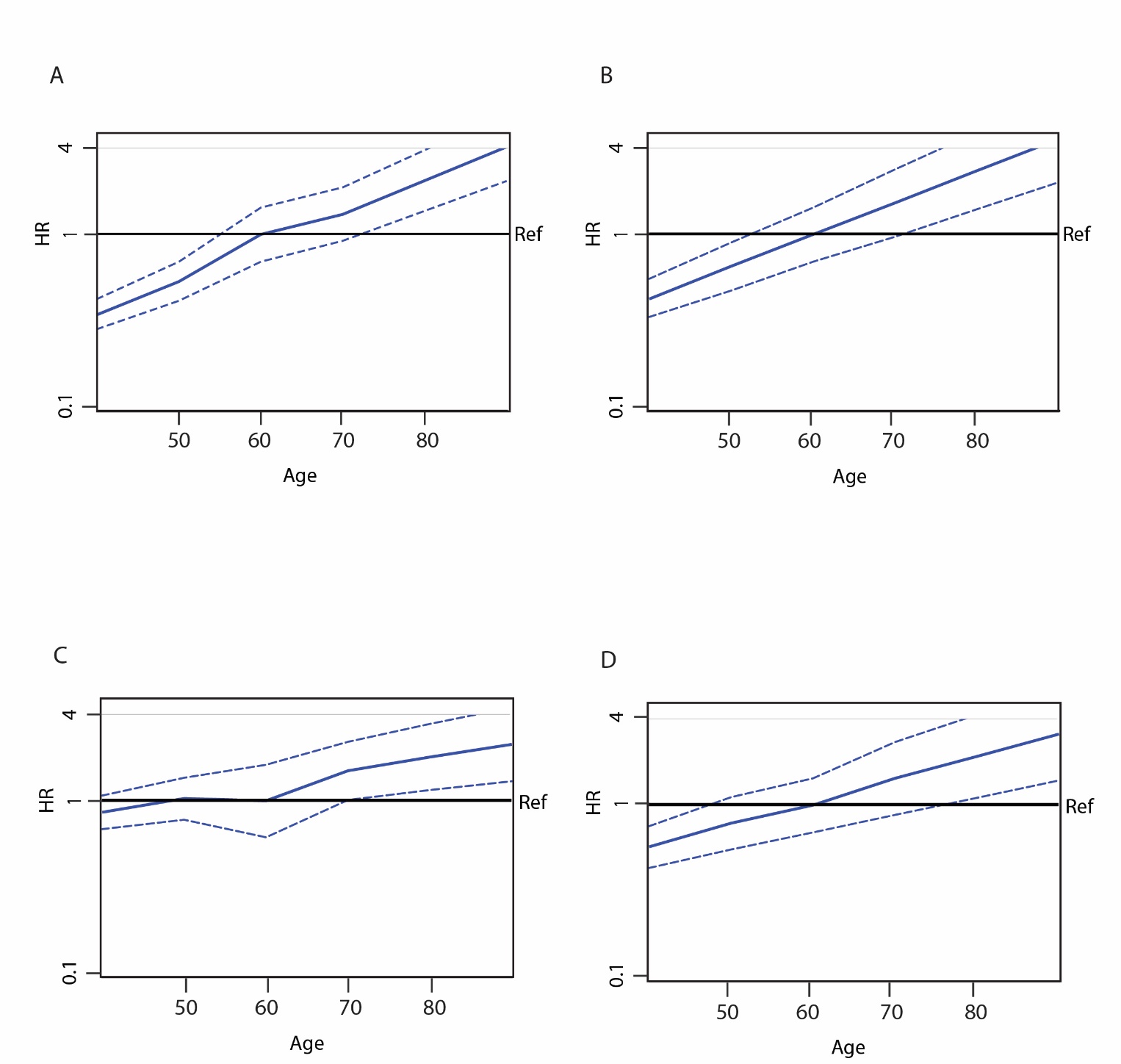

### Supplementary figure 4. Correlation of age with the risk of secondary outcomes using an AutoCI model

A) Correlation of age with the risk of distant metastasis by causal inference. B) Correlation of age with the risk of locoregional recurrence by causal inference. C) Correlation of age with the risk of pelvic recurrence by causal inference. D) Correlation of age with the risk of vaginal recurrence by causal inference. Definition of abbreviations: HR = hazard ratio. Ref = reference line.

### Data Supplement 3. Methods systematic literature search

*Eligibility criteria*

Articles that addressed the impact of calendar age by multivariable analysis on oncological outcomes in women with EC were included. Articles were excluded if: a) the study population consisted (mainly) of stage IV patients, or b) included uterine sarcomas, or c) the study was a case report, (systematic) review, or meta-analysis.

*Search strategy and data selection*

A systematic search was conducted on April 13^th^ 2022 in PubMed. The search string is provided in Data Supplement 4. The identified references were exported to EndNote and screened for eligibility on title and abstract. Full texts were retrieved and eligible articles were selected. In case of >1 article was selected of a single study, the most recent and complete article was used for analysis.

*Data extraction*

Uniform extraction of the following predefined data was performed: first author, country, year of publication, study design, number of patients, disease characteristics (tumour stage, grade, histology, lymphovascular space invasion [LVSI], and molecular class), treatment characteristics (lymphadenectomy, adjuvant treatment), the impact of age on oncological outcomes (vaginal-, pelvic-, and distant recurrences, recurrence-free survival, cancer-specific survival) in multivariable analysis, correction factors and definition of any cut-off for advanced age.

*Data synthesis*

The aforementioned predefined data of all selected articles were aggregated to create an overview of the current knowledge on prognostic impact of age on oncological outcomes. This includes: the applied definition of advanced age (use of any cut-offs), the type of oncological outcome(s), significance of the prognostic impact of age in multivariable analysis. In addition, the quality of the correction for confounding on any of the five established prognostic factors (stage, histotype, grade, LVSI and the molecular classification) was summarized in a score ranging from 0 to 5. Finally, data synthesis was also stratified by study design, to investigate impact of confounding by indication (affecting result of non-randomised studies) on the relation between age and oncological outcomes.

*Overview of search*

The systematic search identified 783 articles. 204 articles were selected for full-text review, of which 108 were included for data synthesis. Results of the search can be found in Supplementary Table 4.

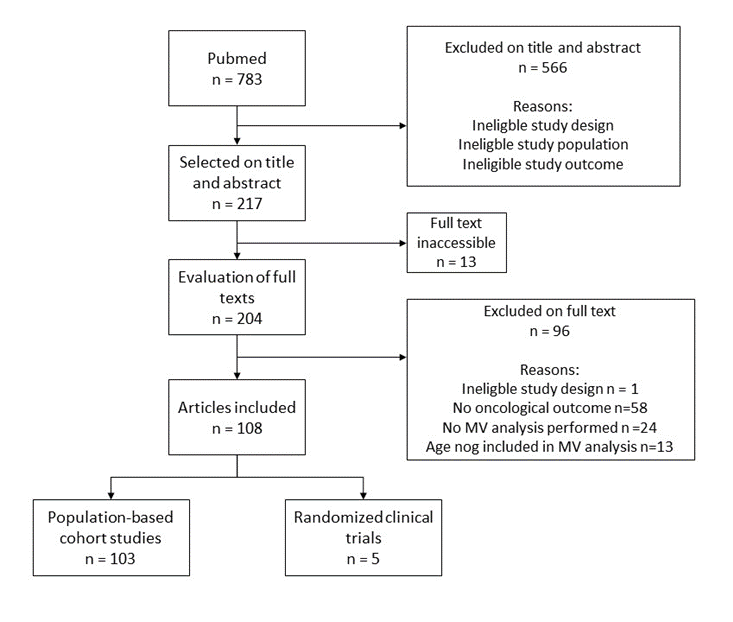

### Data Supplement 4. Search string

("Endometrial Neoplasms"[Mesh:NoExp] OR "Carcinoma, Endometrioid"[Mesh] OR "Endometrial cancer"[tiab] OR "Endometrial carcinoma"[tiab] OR "Uterine cancer"[tiab] NOT ("Endometrial Neoplasms/diagnostic imaging"[Majr] OR "Endometrial Neoplasms/diagnosis"[Majr] OR "Endometrial Neoplasms/metabolism"[Majr] OR "Endometrial Neoplasms/economics"[Majr] OR "Endometrial Neoplasms/immunology"[Majr] OR "Endometrial Neoplasms/genetics"[Majr]))

AND

("Age Factors"[Mesh:NoExp] OR "Age of Onset"[Mesh] OR "Adult"[Mesh] OR "Age"[tiab] OR "Elderly"[tiab])

AND

("Survival"[Mesh] OR "Survival Analysis"[Mesh] OR "Prognosis"[Mesh:NoExp] OR "Treatment Outcome"[Mesh:NoExp] OR "Disease-Free Survival"[Mesh] OR "Progression-Free Survival"[Mesh] OR "Treatment Failure"[Mesh:NoExp] OR "Neoplasm Recurrence, Local"[Mesh] OR "Neoplasm Metastasis"[Mesh:NoExp] OR "Survival Rate"[Mesh] OR "Prognosis"[tiab] OR "Locoregional recurrence"[tiab] OR "Recurrence"[tiab] OR "Overall survival"[tiab])

AND

("Risk Factors"[Mesh:NoExp] OR "Multivariate Analysis"[Mesh:NoExp] OR "Prognostic factor*"[tiab] OR "Risk Factor*"[tiab] OR "Multivariable"[tiab] OR "Multivariate"[tiab] OR "regression"[tiab] OR "Competing risk*"[tiab] OR "*stratif*"[tiab] OR "*standardi*"[tiab] )

NOT

("Case Reports"[Publication Type] OR "Meta-Analysis"[Publication Type] OR "Systematic Review"[Publication Type] OR "Review"[Publication Type] OR "Introductory Journal Article" [Publication Type] OR "Publication Components" [Publication Type] OR "Clinical Conference"[Publication Type] OR "Consensus Development Conference"[Publication Type] OR "Scientific Integrity Review"[Publication Type] OR "Antibodies"[Mesh] OR "Robotic Surgical Procedures"[Mesh] OR "Immunotherapy"[Mesh] OR "immunotherapy"[tiab] OR "Immunohistochemistry"[Mesh] OR "pembrolizumab"[tiab] OR "Immune Checkpoint Inhibitors"[Mesh] OR "DNA"[tiab] OR "RNA"[tiab] OR "pathway"[tiab] OR "signaling"[tiab] OR "Inoperable"[tiab] OR "Irresectable"[tiab] OR "endometriosis"[tiab] OR "adenomyosis"[tiab] OR "Endometriosis"[Mesh] OR "Adenomyosis"[Mesh] OR "Laparoscopy"[Mesh] OR "Laparotomy"[Mesh] OR "Uterine Myomectomy"[Mesh] OR "Fertility"[Mesh] OR "Fertility Preservation"[Mesh] OR "Insulin Resistance"[Mesh] OR "Endometrial Hyperplasia"[Mesh] OR "Ovarian Neoplasms"[Mesh] OR "Male"[Mesh] OR "Neoplasm, Residual"[Mesh] OR "Operative Time"[Mesh] OR "Length of stay"[Mesh] OR "Blood loss, surgical"[Mesh] OR "Blood Transfusion"[Mesh] OR "Cytoreduction Surgical Procedures"[Mesh] OR "Neoplasm Staging"[Majr] OR "Postoperative Nausea and Vomiting"[Mesh] OR "Radiation Injuries"[Mesh] OR "Lymph Node Excision"[Mesh] OR "Venous Thromboembolism"[Mesh] OR "African Americans/genetics"[Mesh] OR "Asians/genetics"[Mesh] OR "Hispanic or Latino/genetics"[Mesh] OR "Whites/genetics"[Mesh] OR "Palliative Care"[Mesh] OR "Time-to-Treatment/statistics and numerical data"[Mesh] OR "Ambulatory Surgical Procedures"[Mesh] OR "Hysterectomy/methods"[Mesh] OR "Peritoneal cytology"[tiab] OR "Poly-ADP-Ribose Binding Proteins"[Mesh] OR "Hysterectomy/adverse effects"[Mesh] OR "Hysterectomy/methods"[Mesh] OR "Peritoneal Cavity"[Mesh] OR "Hysteroscopy"[Mesh] OR "Hysteroscopy"[tiab] OR "Neuroendocrine Tumors"[Mesh] OR "Breast Neoplasms"[Mesh] OR "Colorectal Neoplasms"[Mesh] OR "Uterine Cervical Neoplasms"[Mesh])

### Supplementary Table 5. Results systematic literature search

| **Oncological outcome^1^** | | **Age identified as risk factor^2^** | **Correction for confounding^3^** | | **Definition of advanced age^4^** | |
| --- | --- | --- | --- | --- | --- | --- |
| Disease-specific survival | 28 (21.5%) | 19 (67.9%) | 0 factors | 6 (5.6%) | Continuous | 35 (35.4%) |
| Recurrence-free survival | 84 (64.6%) | 43 (51.2%) | 1 factor | 25 (23.1%) | Ordinal | 12 (12.1%) |
| Distant metastasis | 8 (6.2%) | 2 (25.0%) | 2 factors | 34 (31.5%) | Binary >60 years | 33 (33.3%) |
| Locoregional recurrence | 8 (6.2%) | 5 (62.5%) | 3 factors | 28 (25.9%) | >65 years | 8 (8.1%) |
| Pelvic recurrence | 1 (0.8%) | 1 (100%) | 4 factors | 14 (13.0%) | >70 years | 8 (8.1%) |
| Vaginal recurrence | 1 (0.8%) | 1 (100%) | 5 factors | 1 (0.9%) | >75 years | 3 (3.0%) |

Data synthesis of a systematic literature search on the prognostic impact of age on oncological outcomes in women with stage I-III endometrial cancer. 108 studies were included in the analysis with a total of 196,862 patients. 1) Some studies reported on more than one outcome. 2) Indicates whether age was identified as a risk factor for worse oncological outcomes by multivariable analysis. 3) The number of the five established risk factors that was corrected for in multivariable analysis (stage, histotype, grade, lymphovascular space invasion and molecular class). 4) Nine of 108 studies had a different definition of age, namely they used the median age of their data set as cut-off, or random values (40, 50, 55, 58, 59, 68).
